# Analysis of key factors of a SARS-CoV-2 vaccination program: A mathematical modeling approach

**DOI:** 10.1101/2021.02.19.21252095

**Authors:** David Martínez-Rodríguez, Gilberto Gonzalez-Parra, Rafael-J. Villanueva-Micó

## Abstract

The administration of vaccines against the coronavirus disease 2019 (COVID-19) just started in early December of 2020. Currently, there are only a few approved vaccines, each with different efficacies and mechanisms of action. Moreover, vaccination programs in different regions may vary due to differences in implementation, for instance, simply the availability of the vaccine. In this article, we study the impact of the pace of vaccination and the intrinsic efficacy of the vaccine on prevalence, hospitalizations, and deaths related to the SARS-CoV-2 virus. Then we study different potential future scenarios regarding the burden of the COVID-19 pandemic in the near future. We construct a compartmental mathematical model and use computational methodologies to study these different scenarios. Thus, we are able identify some key factors to reach the aims of the vaccination programs. We use some metrics related to the outcomes of the COVID-19 pandemic in order to assess the impact of the efficacy of the vaccine and the pace of the vaccine inoculation. We found that both factors have a high impact on the outcomes. However, the rate of vaccine administration has a higher impact in reducing the burden of the COVID-19 pandemic. This result shows that health institutions need to focus in increasing the vaccine inoculation pace and create awareness in the population about the importance of COVID-19 vaccines.

## 1 Introduction

The world is facing the COVID-19 pandemic and just in the middle of December of 2020 the first vaccines are being given to a few people [42, 128, 78, 119, 103, 129, 130, 115]. This pandemic has caused more than 80 million confirmed cases and more than 1.7 million deaths [30, 136].

The SARS-CoV-2 virus causes an illness called COVID-19 that can result in severe pneumonia and death [81, 102]. The complex process of the SARS-CoV-2 spread involves several factors that are currently not very well understood [84, 112, 139, 102, 31, 32]. There are several aspects that impact the spread of the virus in the human population, such as, social behavior, age, weather variables, mutation of the virus, and immunocompetence [152, 111, 147]. Other factors may affect the spread of the SARS-CoV-2 virus but still be unknown. Regarding mutations, SARS-CoV-2 could acquire mutations with fitness advantages and immunological resistance [71]. Therefore, studying evolutionary transitions is important to ensure effectiveness of the vaccines and immunotherapeutic interventions [151, 50, 104, 157, 71]. It has been stateded that not only is the efficacy of the vaccine important, but whether a vaccine reduces infection and transmission as well as disease progression [69].

The genomic analysis suggested that the base sequence of SARS-CoV-2 is almost 80% similar to that of SARS-CoV. Further, both these viruses bind to same host cell receptor ACE-2 [6]. However, the development of vaccines for this novel SARS-CoV-2 virus took nearly a year. There are now more than 80 vaccines in development [3, 16, 69, 71, 149, 105, 135]. Vaccination programs have recently begun (December) in a few countries, and there are many uncertainties regarding the optimal implementation of these vaccination programs and the probable outcomes [1, 69, 137]. Therefore, studying the COVID-19 vaccination programs is of paramount importance. Effective vaccination helps tackle the transmission of the SARS-CoV-2 virus in the population [9, 29, 69, 72, 137, 145].

Mathematical models, statistical analyses and computational techniques are very useful tools to study different processes, including testing hypotheses and understanding how factors affect the processes. For infectious disease processes, mathematical models can be used to perform *in silico* simulations of different potential scenarios, vaccination programs, and test different strategies to slow down epidemics [92, 12, 73, 40, 2, 20, 114, 45, 5, 46]. The outcomes of the complex infectious disease processes under different scenarios are generally impossible to predict without mathematical models and computational techniques. In some cases, results of the simulations can be counter-intuitive and very interesting from a predictive point of view.

There are many articles related to the use of mathematical modeling in combination with computational and statistical techniques to study the spread of the SARS-CoV-2 virus [31, 73, 126, 40, 130, 74, 155]. Some mathematical models used the SIR (Susceptible-Infected-Recovered) mathematical model [107, 8, 110]. Previous studies have used susceptible-exposed-infected-recovered (SEIR) type models [130, 61, 153]. Other mathematical models use a curve fitting of some particular growth model to the data, and also artificial intelligence techniques have been considered for fitting models to real data related to COVID-19 [41, 118, 80].

The main advantage of mathematical models is that many different simulations can be done and this allows us to study the main driving factors of pandemics under a variety of complex scenarios [92, 12, 73, 40, 20, 114, 45, 5, 46]. However, many forecasts related to the COVID-19 pandemic disagree with each other due to many related uncertainties in key characteristics of the SARS-CoV-2 virus [117, 59, 38, 64, 125, 133, 73, 126, 40, 130, 74, 155]. Moreover, currently we are facing new strains due to mutations of the virus, which has raised questions about the efficacy of the vaccines against the mutations of the SARS-CoV-2 virus. Recently, it has been found that the SARS-CoV-2 is mutating and its transmission is more efficient [151, 98, 33, 77]. There is a growing literature about mutations of the SARS-CoV-2, but it is not clear what further mutations could occur in the near future [151, 50, 104, 157, 71].

A new vaccine campaign against the SARS-CoV-2 virus began in December of 2020 in the United States and other countries. Currently, there are only a few approved vaccines with different efficacies and mechanisms of action. Our principal aim in this article is to study the impact of the pace of vaccination and the efficacy of the vaccine on the outcome or dynamics of the incidence, prevalence, and deaths related to the SARS-CoV-2 virus [58, 70, 79, 82, 101]. This will help us to explain different potential patterns in different countries related to their vaccination programs [83, 98, 112, 51]. Despite the huge health crisis caused by the spread of the SARS-CoV-2 virus around the world, there are few studies related to the prediction of feasible scenarios in the year 2021 [49, 87, 91, 99]. In [91] the authors used a SEIR mathematical model based on differential equations to study the situation in South-Africa with respect to the number of reported cases of COVID-19. They found that a vaccine with 70% efficacy had the capacity to contain the COVID-19 outbreak but only at a very high vaccination coverage of 94.44%. In [49], the authors proposed a distribution of vaccines in time and space, which sequentially prioritizes regions with the most new cases of infection in a given time period. They used a SEIR type model that includes an extra class for mild infected individuals and with spatio-temporal effects. They found that, for a locally well-mixed population, the proposed strategy strongly reduces the number of deaths. In [87], the authors implemented a SEIR type model that includes pre-symptomatic, asymptomatic and the entire equivalent vaccinated classes. They studied the impact of different vaccination coverages, efficacy and reduction of symptoms on different metrics such as deaths, and ICU hospitalizations. The outcomes of these studies are important because they help to understand better the impact of vaccination programs, and generate optimal actions to diminish the spread of the SARS-CoV-2 virus [73, 126, 40, 55, 87, 152, 111, 147]. One factor that brings uncertainty to the outcomes and that must be taken into account is the fact that vaccines are under threat in different places and there are antivaccine movements that have gained traction with some people [1, 27].

In this article, we construct a compartmental mathematical model and use computational methodologies to study different scenarios. In particular, we include the asymptomatic carriers of the virus, who are nevertheless able to spread the virus. It has been mentioned that asymptomatic people are in some way a key contributor in the spread of the SARS-CoV-2 virus, and are real threat for the control policies [7, 62, 90, 100, 124, 43, 68, 52, 132, 31, 62]. For instance, it has been found that quantitative SARS-CoV-2 viral loads were similarly high for infected individuals with symptoms, pre-symptomatic, or asymptomatic. Moreover, it has been found 6 to 24 times more estimated infections per site with seroprevalence than with coronavirus disease 2019 (COVID-19) case report data [56]. Thus, we construct a mathematical model taking into account asymptomatic people, which have been missed in other studies. In addition, we will study scenarios with different SARS-CoV-2 virus transmission rates, which results in different effective reproduction numbers *R*_*t*_ of COVID-19 [75, 85, 14, 25].

## 2 Materials and Methods

### 2.1 Mathematical Model

We constructed a compartmental model based on differential equations that includes individuals in the susceptible, latent, infected, asymptomatic, and hospitalized stages. The mathematical model considers transitions of individuals through the aforementioned stages depending on the COVID-19 progression. In addition to the previous stages, the model incorporates vaccinated individuals that might be in analogous stages such as susceptible or asymptomatic vaccinated. Thus, in some way we can classify individuals in two disjoint groups: unvaccinated and vaccinated. We assume that unvaccinated individuals in the susceptible, latent, and asymptomatic compartments can receive the vaccine against the SARS-CoV-2 virus. On the other hand, we assume that symptomatic, recovered and hospitalized unvaccinated individuals do not receive the vaccine. The individuals can transit from the unvaccinated susceptible class to vaccinated susceptible if they get the vaccine. In an analogous way, the latent and asymptomatic unvaccinated individuals can move to the respective vaccinated compartment. It is important to mention that the model incorporates the type of vaccine that diminishes the progression to the COVID-19 disease [87, 99]. Individuals in the latent stage are not yet infectious. The individuals remain in the latent stage for a certain time which is chosen from an exponential distribution with mean time *α*. The individuals then transit into the infective symptomatic or asymptomatic stages, where they are able to spread the SARS-CoV-2 virus to other individuals. They stay in the infectious stage for a time chosen from an exponential distribution with mean time *γ*. After that, individuals in the asymptomatic stage move to the recovered stage. However, individuals in the infective symptomatic stage can move to the recovered or to the hospitalized stages, depending on the level of disease progression. Even though we assume exponential distributions, the Erlang distributions are more realistic but at the expense of more complex models and more parameters [63, 48, 116, 47, 38, 140]. Thus, many studies assume exponential distributions to avoid greater complexity in the models and in the analysis. However, in some cases exponential distributions are not far from reality. We have found that the length of stay in the hospital is not far from an exponential distribution [37]. Finally, hospitalized individuals can die due the COVID-19 disease [37, 39, 40, 156]. This last metric (or outcome) is of paramount importance [40, 138, 142, 150].

We use a mathematical model that is similar to a *SEIR*-type epidemiological model to explain the dynamics of COVID-19 spread on the human population under a vaccination program. This model has parameters that can be varied in order to study different possible scenarios. For instance, the pace of vaccination and efficacy of the vaccine can be modified. This is important since it is known that the efficacy of vaccines varies and they have different underlying mechanisms of action [69, 72, 58, 79, 9]. Moreover, different countries and regions would apply the vaccines at different rates due to a variety of factors such as availability and resources [1, 35, 69, 79, 27].

The constructed mathematical model based on differential equations is given by

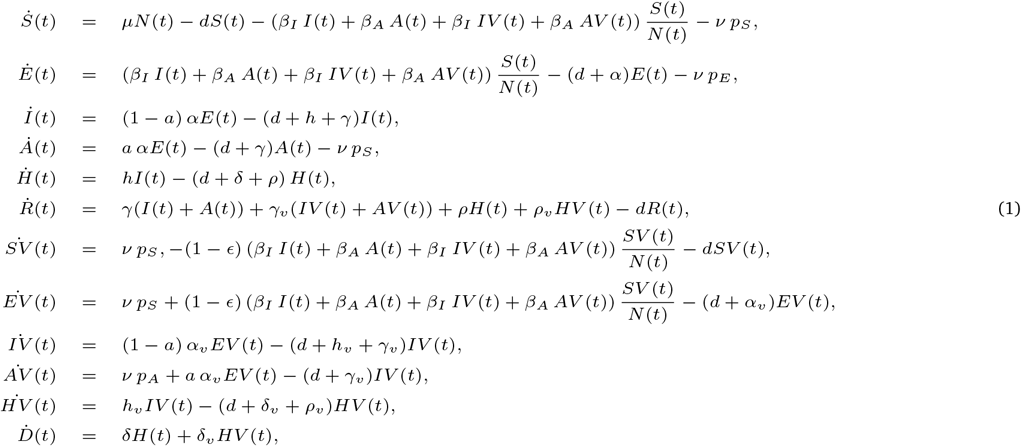

where *S*(*t*) denotes the number of susceptible individuals. When a susceptible and an infectious individual come into infectious contact, the susceptible individual contracts the disease and transitions to the latent compartment *E*(*t*). Individuals in compartment *E*(*t*) are infected (carry the virus) but cannot spread the virus. Compartment *I*(*t*) represents individuals who have been infected and show symptoms. These individuals are capable of infecting susceptible individuals after being in the *E*(*t*) subpopulation. The subpopulation *A*(*t*) represents the number of individuals who have been infected but are asymptomatic. These individuals are capable of infecting susceptible individuals after being in the *E*(*t*) subpopulation. The variable *H*(*t*) denotes the number of hospitalized individuals at time *t*. The compartment *D*(*t*) represents the number of deaths due to the SARS-CoV-2 virus from the beginning of the simulation period. Similarly, *SV* (*t*), *EV* (*t*), *IV* (*t*), *AV* (*t*) and *HV* (*t*) denote the analogous vaccinated population at time *t*. Individuals in the *S*(*t*), *E*(*t*) and *A*(*t*) classes are vaccinated with rates *ν p*_*S*_, *ν p*_*E*_ and *ν p*_*A*_ respectively. These proportions are related to the their respective size populations. The model assumes that people in states *E*(*t*), *EV* (*t*), *HV* (*t*), *H*(*t*), and *R*(*t*) do not transmit the infection.

In this model we consider that COVID-19 confers immunity after recovery (currently assumed but not confirmed), and assume that when an individual is in the latent and latent vaccinated stages the virus cannot be transmitted. In addition, we consider that once the individuals receive the vaccine inoculation they transit to the vaccination compartments. The model assumes one vaccine inoculation to cause the individual to transit to one of the vaccinated compartments. The model also considers that hospitalized individuals are not able to transmit the SARS-CoV-2 virus. This assumption is arguable, but we assume the conditions in the hospitals are safe regarding the transmission of the SARS-CoV-2 virus. We also assume that individuals in the susceptible, latent, and asymptomatic compartments are those that can be vaccinated. This assumption might sound arguable, but recently it has been mentioned that a nurse tested positive for COVID-19 more than a week after receiving Pfizer Inc’s vaccine [113]. One hypothesis to explain this observation is that the nurse was in the latent stage before being inoculated.

### 2.2 Parameter values

In this work we are interested in the impact of the vaccination rate and the efficacy of the vaccines on the infected, hospitalized, and death cases. We assume that the rates of virus transmission in asymptomatic and symptomatic individuals are constant from the beginning of the period of study i.e. when the vaccination program starts. This implicitly assumes that people would not change behavior (on average) until the vaccination program is well advanced. This is a credible assumption in the USA, and previous physical and social behavior changes can be included in the transmissibility. Many health policies and guidelines would have been implemented before the vaccination program started. In some cases it is more realistic to include time-varying transmissibility, which has been used to study other infectious diseases and in particular one closely related to the SARS-CoV-2 virus [131, 76, 73, 67]. This latter approach is more troublesome to implement since it is necessary to estimate a time-varying parameter, and identifiability issues thus arise. Even with accurate data from the past it is difficult to estimate a time-varying transmission. Moreover, in this study we can not predict how the behavior of individuals might change in the future. Thus, we take an approximation and a conservative assumption that the transmissibility would not change during the beginning of the vaccination program.

We assume that the parameters related to the COVID-19 disease progression are the same for vaccinated and unvaccinated. In addition, we consider that vaccinated individuals are not able to get the disease unless the vaccine was not effective. This aspect is not clear in the relevant scientific literature due to the different types of COVID-19 vaccines [69, 72, 58, 79, 9]. In addition, some studies have indicated that the antibody titers may decline over time in patients recovered from COVID-19, particularly in those who were asymptomatic [143]. However, we do not consider that recovered individuals can return to the susceptible stage. One reason for this is that further studies are needed to check how long the immunity lasts, and furthermore, the time horizon of this study is less than 16 months. We also consider that for this period the immunity provided by the vaccines does not diminish.

Reasons for doubting this undiminished immunity come from studies on the SARS virus. For example, in a study of 56 patients recovered from SARS it was found that the neutralizing and IgG antibodies quickly declined after 16 months and continued to decline further to a very low level after 3 years [143]. Moreover, trials of SARS vaccines also suggest that the neutralizing antibody responses may decline over time [143]. On the other hand, it is unclear whether vaccine induced antibody levels could persist and, if not, whether the long-lasting memory T cells could affect susceptibility and pathogenesis of SARS-CoV-2 infection [143, 19].

It has been mentioned that US federal officials hoped for twenty million people to get their first of two required shots by the end of 2020. However, they recently changed that goal and just over 1 million doses of vaccines had been administered (Dec. 26th 2020) [134]. Therefore, we assume as a lower bound rate *ν* for the inoculation of the vaccine a value of one million per week. This rate can be increased since it is expected that the process of the vaccine administration will be improved. However, this value of the parameter is subject to variation due to the reluctance of some people to vaccinate for because of doubtfulness about the preliminary tests of efficacy to pressure from anti-vaccination movements [27].

For the death rate of hospitalized individuals we use a variety of data from the scientific literature [40, 66, 148, 91, 99]. We used the weighted average of the probability of dying for severe and critical cases (ICU), and in addition we took into account the average length of stay in the hospital [99]. We varied in a reasonable way the death rate in order to take into account the possible uncertainty in the data.

For the asymptomatic cases and proportions we also relied on data from the scientific literature [21, 88, 40, 68, 90, 97, 96, 154]. However, the discrepancies in the relevant data are great. We chose as a conservative starting point that the percentage of infections that are asymptomatic is 50% [30]. However, for the numerical simulations we additionally considered a percentage of 40% [30, 96].

For the *β*_*s*_ parameters we assume values in the range of [0.1 − 0.5], which are values found in some studies. Currently, in USA there are several non-pharmaceutical interventions. We also assume for the numerical simulations that *β*_*A*_ *≤β*_*I*_. This assumption is based on the uncertainty in these values as well as results from the literature that the infectiousness of asymptomatic carriers is similar or smaller to the symptomatic [21, 88, 40, 68, 121, 154]. One interesting article found that asymptomatic carriers have a higher viral load, and, taking into account that asymptomatic carriers might have more physical contacts, it is possible to assume that *β*_*A*_ *≥ β*_*I*_ [54].

### 2.3 Initial conditions for the scenarios

For the initial conditions we assume the particular situation of the USA since is one of the first countries that started a vaccination program [134, 106]. We rely on data from the scientific literature and demographics of the USA. As expected, there are some uncertainties related to data of the COVID-19 pandemic and which is usual in many epidemics. For instance, the infected reported cases have uncertainties due to many factors such sensitivity and specificity of COVID-19 tests [10, 127]. Moreover, asymptomatic cases represent a great uncertainty [31, 43, 52, 62, 90, 100, 124, 132, 7]. Taking into account these uncertainties, we set the initial conditions presented in Table 2. The total initial population *N* (0) is taken from the current USA population [15]. The birth and death rates are taken from the official website of the CDC in USA [144, 86]. All the initial vaccinated subpopulations, are set to zero since the simulations are performed at the beginning of the vaccination program. Two key initial subpopulations are those corresponding to the infected and asymptomatic, since they affect the initial dynamics of the COVID-19 pandemic under the vaccination program. We took the seven day average of the infected reported cases and then multiplied by seven days (assumed infectiousness period) and by 0.8 to obtain the initial number of symptomatic cases (assumption of the proportion of symptomatic cases in the reported cases) [17, 4, 89, 123, 23, 154, 65, 122, 44]. The percentage of asymptomatic cases in the official statistics varies for each country. In some countries it may be close to zero, since no random tests are performed. However, the detection of asymptomatic infections is possible in the case of the USA in situations in which testing is mandatory (as in some universities) or random. We approximated this value by relying on data from different studies [17, 4, 89, 123, 23, 154, 65, 122, 44]. However, in our simulations we varied the parameter through reasonable values. This variability only affects the initial conditions of some of the populations.

In the reported cases we have a subpopulation of asymptomatic cases since the testing programs take into account the entire population. We take the initial subpopulation of asymptomatic carriers as equal to the symptomatic one. This implicitly assumes that the percentage of asymptomatic cases is 50%. We use this value based on information from the CDC official website, even though it is mentioned that there is uncertainty in this percentage. We found that there is a large uncertainty in scientific literature regarding this percentage [30, 31, 43, 52, 62, 90, 100, 124, 132, 7]. Therefore, we vary it in order to deal with its uncertainty. For the initial latent subpopulation we take into account that the latent period is around 5.2 days and the latent stage includes individuals who will become either asymptomatic or symptomatic [40]. For the initial hospitalized subpopulation we take into account that hospitalized individuals spend an average of 10.4 days in the hospital and that around 4% of the symptomatic infected transit to the hospitalization stage [40, 78, 60]. For the recovered COVID-19 cases we take into account the current total of reported infected cases and the fact that a subset of the asymptomatic cases are not reported. In addition, we notice that we need to subtract the current number of infected and asymptomatic cases. This approximation gives us a plausible number of recovered cases that exceeds the reported recovered cases (*≈* 11 millions) [30, 136]. Finally, we use for the initial susceptible subpopulation the fact that initially there are not vaccinated individuals and therefore *S*(0) = *—N* (0) *−E*(0) *−I*(0) *−A*(0) *−R*(0) *− H*(0). In Table 2, we present the initial conditions for the subpopulations.

## 3 Results

In this section, we perform numerical simulations of the mathematical model (1) to analyze the impact of the vaccination rate and the efficacy of the vaccine on the dynamics of the COVID-19 pandemic. We use the parameter values of Table 1 and the initial conditions given in Table 2. We vary the values of the vaccination rate, the efficacy of the vaccine and the transmission rates in order to include a variety of scenarios that take into account the uncertainty in the aforementioned factors. We introduce some important metrics related to the outcomes of the COVID-19 pandemic in order to assess the impact of the inoculation rate and the efficacy of the vaccine.

**Table 1:** Mean values of parameters used to perform numerical simulations of the different scenarios.

| Parameter | Symbol | Value |
| --- | --- | --- |
| Incubation period | $\alpha^{-1}$ | 5.2 days [78, 131] |
| Infectious period | $\gamma^{-1}$ | 7 days [78] |
| Hospitalization rate | $h^{-1}$ | 3.5 days × 0.04 [40, 78, 60] |
| Hospitalization period | $\rho^{-1}$ | 10.4 days [40, 78, 60] |
| Death rate (hospitalized) | $\delta^{-1}$ | 10.4 days × 0.103 [109, 99] |
| Probability of being asymptomatic | $a$ | 0.5 [30, 96] |
| Birth rate | $\mu^{-1}$ | 0.00003178 days [15] |
| Death rate | $d^{-1}$ | 0.00002378 days [15] |

**Table 2:** Initial conditions assumed for the different subpopulations using the USA current situation (Early-December).

| Parameter | Symbol | Value |
| --- | --- | --- |
| Latent | $E(0)$ | 1, 782, 857 |
| Infected (symptomatic) | $I(0)$ | 1, 200, 000 |
| Asymptomatic | $A(0)$ | 1, 200, 000 |
| Hospitalized | $H(0)$ | 71, 552 |
| Susceptible | $S(0)$ | 309, 974, 354 |
| Recovered | $R(0)$ | 16, 462, 937 |
| Total population | $N(0)$ | 330, 705, 643 |

### 3.1 Vaccination rate, efficacy and transmission rate scenarios

Here we present the results of the numerical simulations for different scenarios varying the inoculation rate, efficacy of the vaccine, percentage of infections that are asymptomatic, and the transmission rates. We consider two different plausible efficacies for the vaccines. We set the efficacy (*E*) to 80% and 94%. These values were chosen based on some results of vaccine trials and the current approved vaccines [3, 16, 69, 134, 149, 105, 135]. We could simulate scenarios with lower efficacies if we desire and based on the fact that the FDA established a minimum efficacy threshold of at least 50% [58, 99]. We also vary the inoculation rate (vaccination pace) to test different potential vaccination program scenarios [134, 106]. It is important to remark that despite the plans that health institutions make regarding vaccination, there are uncertainties present in the logistics [134, 95, 141, 106]. For instance, currently there is a significant delay in coronavirus vaccinations while hospitalizations continue to set records in the USA [106]. Therefore, here we considered two different plausible inoculation rates based on the current situation. Specifically, we chose vaccination rates of two and four millions per week. It is important to mention that even though these rates might not be 100% accurate, this approach helps to elucidate the impact of the inoculation rate on the main outcomes of the COVID-19 pandemic under a vaccination program.

Regarding the values of the SARS-CoV-2 virus transmission rate that plays an important role in the value of the effective reproduction number ℛ_*t*_, we chose two different values. These two values of the SARS-CoV-2 virus transmission rate between humans correspond to two different reproduction numbers ℛ_0_. Thus, we can relate them using the following equation 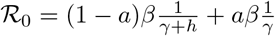. It is important to remark that the effective reproduction number ℛ_*t*_ varies over the time, and several methods have been proposed to compute it [24, 146, 120, 94, 75, 26, 131]. For instance, under certain conditions ℛ_*t*_ = ℛ_0_*S*(*t*)*/N*, which relates the value of the virus transmissibility *β* to the effective reproduction number [146]. It is important to remark that independently of the method that is used to compute the effective reproduction number ℛ_*t*_ all of them show that it depends explicitly or implicitly on the value of the transmission rates (*β*_*s*_). Therefore varying these rates implies a variation in the basic reproduction number ℛ_0_ and on the effective reproduction number ℛ_*t*_. Thus, we are considering different scenarios regarding the risk of becoming infected by the SARS-CoV-2.

The aim is to test the impact of the inoculation rate and efficacy of the vaccine under two different SARS-CoV-2 virus transmission rate scenarios. As it has been mentioned before, there are a lot of uncertainties in the transmission rates for different regions and they vary over the time depending on official and unofficial non-pharmaceutical interventions [40, 55, 85, 93, 120, 25, 75]. However, the approach used here allows us to understand the impact of inoculation rate and vaccine efficacy under two different transmission of the SARS-CoV-2 virus environments. Then results can be extrapolated to other environment settings.

### 3.2 Numerical simulation of scenarios

Here we present the results of the numerical simulations for different scenarios varying the inoculation rate and efficacy, under low transmission rate scenarios. Table 3 shows the peak of the number of infected (symptomatic plus asymptomatic) and hospitalized people for the different vaccine efficacies and inoculation rates. In addition, the number of deaths and recovered cases at the end of the simulation period of 365 days is shown. In this Table we considered several scenarios with two different transmission rates for *β*_*I*_, assumed that the infectiousness of the asymptomatic individuals is the same as the symptomatic (*β*_*A*_ = *β*_*I*_), and that the percentage of infections that are asymptomatic is 50% (*a* = 0.5). It can be seen that the impact of the vaccination rate is greater than the efficacy of the vaccine. For instance, observing the first row of Table 3 it is seen that the peak of the number of infected people is 5,765,525 under a scenario with transmission rate of *β*_*I*_ = 0.2, vaccine efficacy of *E* = 94% and a vaccination rate of two millions per week. Additionally, it is seen that this peak becomes 5,864,871 if the vaccine efficacy decreases to 80%. However, the peak of the number of infected people is 4,791,577, if under the same scenario we change the vaccination rate to four million per week. Thus, it can be seen than the impact of the vaccination rate in the number of infected people is larger than that from vaccine efficacy. We performed additional numerical simulations varying all the parameters in reasonable ranges, and the impact of the vaccination rate was always larger than the vaccine efficacy. In Figure 2, we show the peak of the number of infected and hospitalized individuals for a wide range of different vaccine efficacies and inoculation rates. In addition, the number of deaths and recovered cases are shown. The variation of all these outcomes is larger when the vaccination rate is varied. The results in Table 3 make sense since the effect of the vaccination rate and vaccine efficacy reduce the number of infected, hospitalized and deaths.

**Table 3:** Impact of the inoculation rate (Vac.) and the efficacy (ϵ) of the vaccine on the peak of the infected (I) and hospitalized (H) subpopulations. In addition, on the deaths (D) and recovered (R) cases. In these scenarios, the infectiousness of the asymptomatic individuals relative to symptomatic is 100% (β_A_ = β_I_), and the percentage of infections that are asymptomatic is 50%.

| Pop | Vac.<br>Eff. ($\epsilon$)<br>Trans. ($\beta_I$) | 2 millions | | 4 millions | |
| --- | --- | --- | --- | --- | --- |
|  |  | 94% | 80% | 94% | 80% |
| I | 0.2 | 5.765256e+06 | 5.864871e+06 | 4.791577e+06 | 4.977115e+06 |
|  | 0.22 | 9.002534e+06 | 9.153061e+06 | 7.432046e+06 | 7.762321e+06 |
| H | 0.2 | 2.946735e+05 | 2.998006e+05 | 2.445378e+05 | 2.541361e+05 |
|  | 0.22 | 4.566316e+05 | 4.643245e+05 | 3.768272e+05 | 3.937063e+05 |
| R | 0.2 | 1.315812e+08 | 1.347213e+08 | 1.018598e+08 | 1.079459e+08 |
|  | 0.22 | 1.649504e+08 | 1.681808e+08 | 1.331096e+08 | 1.404869e+08 |
| D | 0.2 | 4.089271e+05 | 4.198674e+05 | 3.052741e+05 | 3.265081e+05 |
|  | 0.22 | 5.255777e+05 | 5.368480e+05 | 4.144543e+05 | 4.402026e+05 |

**Figure 1:**
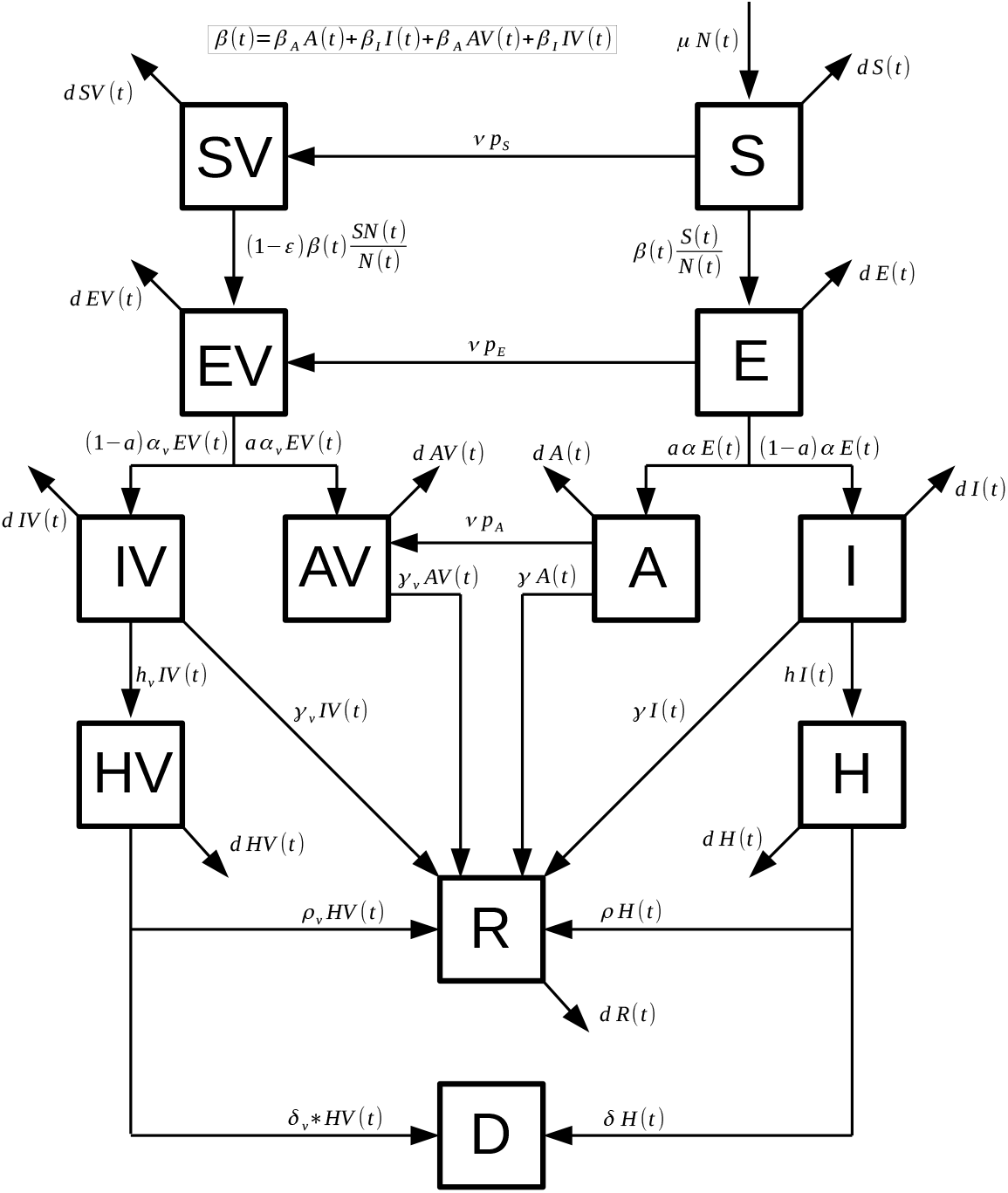
Diagram for the COVID-19 mathematical model (1). The boxes represent the subpopulation and the arrows the transition between the subpopulations. Arrows are labeled by their corresponding model parameters.

**Figure 2:**
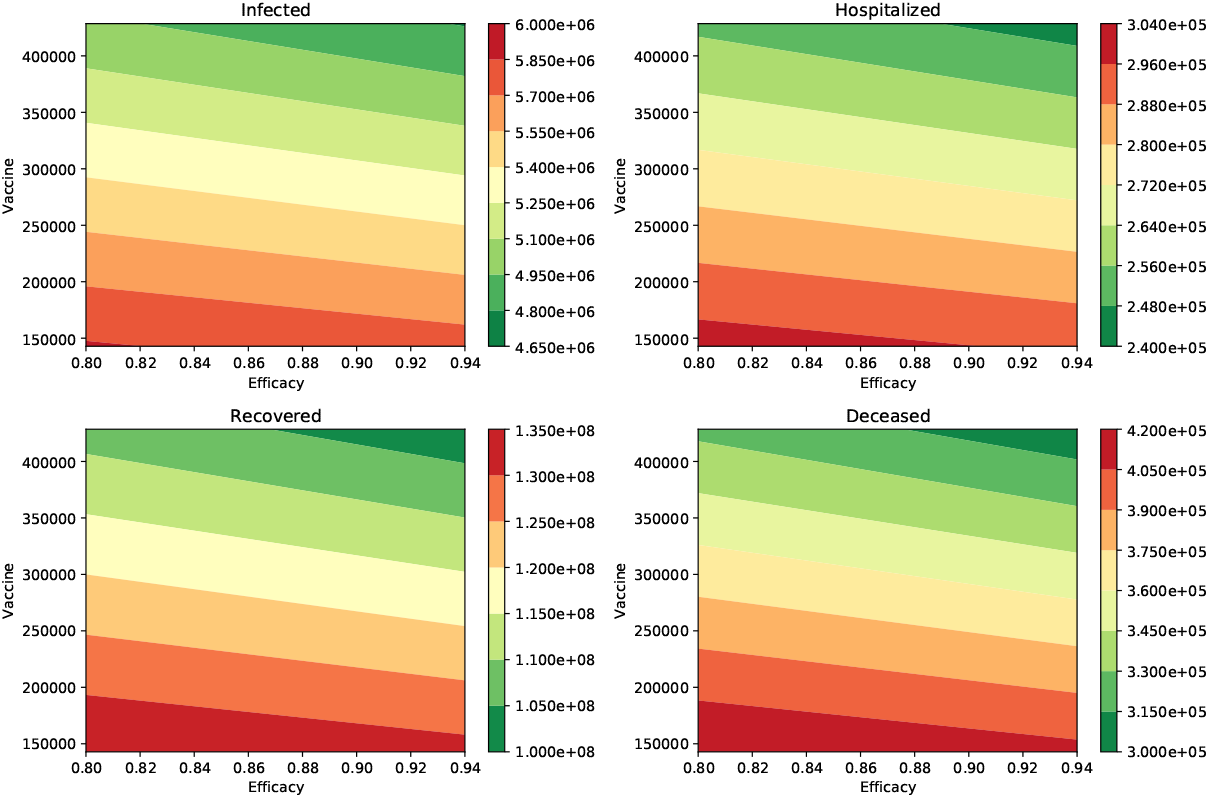
Impact of the inoculation rate and efficacy of the vaccine on the peak of the infected (I) and hospitalized (H) subpopulations. In addition, on the deaths (D) and recovered (R) cases. In these scenarios, the infectiousness of the asymptomatic individuals relative to symptomatic is 100% (β_A_ = β_I_), and the percentage of infections that are asymptomatic is 50%. In all these scenarios the transmission rate is β = 0.2.

We present additional Tables for different scenarios varying the transmission of the asymptomatic carriers and the proportion of asymptomatic individuals. The Table 4 shows the same outcomes that we mentioned above, but we consider now that the infectiousness of the asymptomatic individuals relative to symptomatic is 75%. The impact of the vaccination rate is greater than the efficacy of the vaccine. The first row of Table 4 shows that the peak of the number of infected people is 2,932,727 when transmission rate is *β*_*I*_ = 0.2, the vaccine efficacy is *E* = 94% and the vaccination rate is two millions per week. This metric is just 2,954,850 if the vaccine efficacy decreases to 80%. This is a small change if we compare it to that when the rate of vaccination is increased to four million per week. Thus, this scenario also supports the importance of a high vaccination rate. Figure 3 shows the different outcomes for a wide range of different vaccine efficacies and inoculation rates. The variation of all these outcomes is larger when the vaccination rate is varied.

**Table 4:** Impact of the inoculation rate (Vac.) and the efficacy (ϵ) of the vaccine on the peak of the infected (I) and hospitalized (H) subpopulations. In addition, on the deaths (D) and recovered (R) cases. In these scenarios, the infectiousness of the asymptomatic individuals relative to symptomatic is 75% (β_A_ = β_I_), and the percentage of infections that are asymptomatic is 50%.

| Pop | Vac.<br>Eff<br>Trans | 2 millions |  | 4 millions |  |
| --- | --- | --- | --- | --- | --- |
|  |  | 94% | 80% | 94% | 80% |
| I | 0.2 | 2.932727e+06 | 2.954850e+06 | 2.741826e+06 | 2.773311e+06 |
|  | 0.22 | 4.630903e+06 | 4.704678e+06 | 3.938121e+06 | 4.063971e+06 |
| H | 0.2 | 1.508243e+05 | 1.519984e+05 | 1.404651e+05 | 1.422230e+05 |
|  | 0.22 | 2.372996e+05 | 2.411171e+05 | 2.013844e+05 | 2.079448e+05 |
| R | 0.2 | 8.310916e+07 | 8.523820e+07 | 6.496555e+07 | 6.810785e+07 |
|  | 0.22 | 1.157629e+08 | 1.187113e+08 | 8.874861e+07 | 9.395817e+07 |
| D | 0.2 | 2.395987e+05 | 2.469869e+05 | 1.764218e+05 | 1.873815e+05 |
|  | 0.22 | 3.536476e+05 | 3.639092e+05 | 2.594795e+05 | 2.776527e+05 |

**Figure 3:**
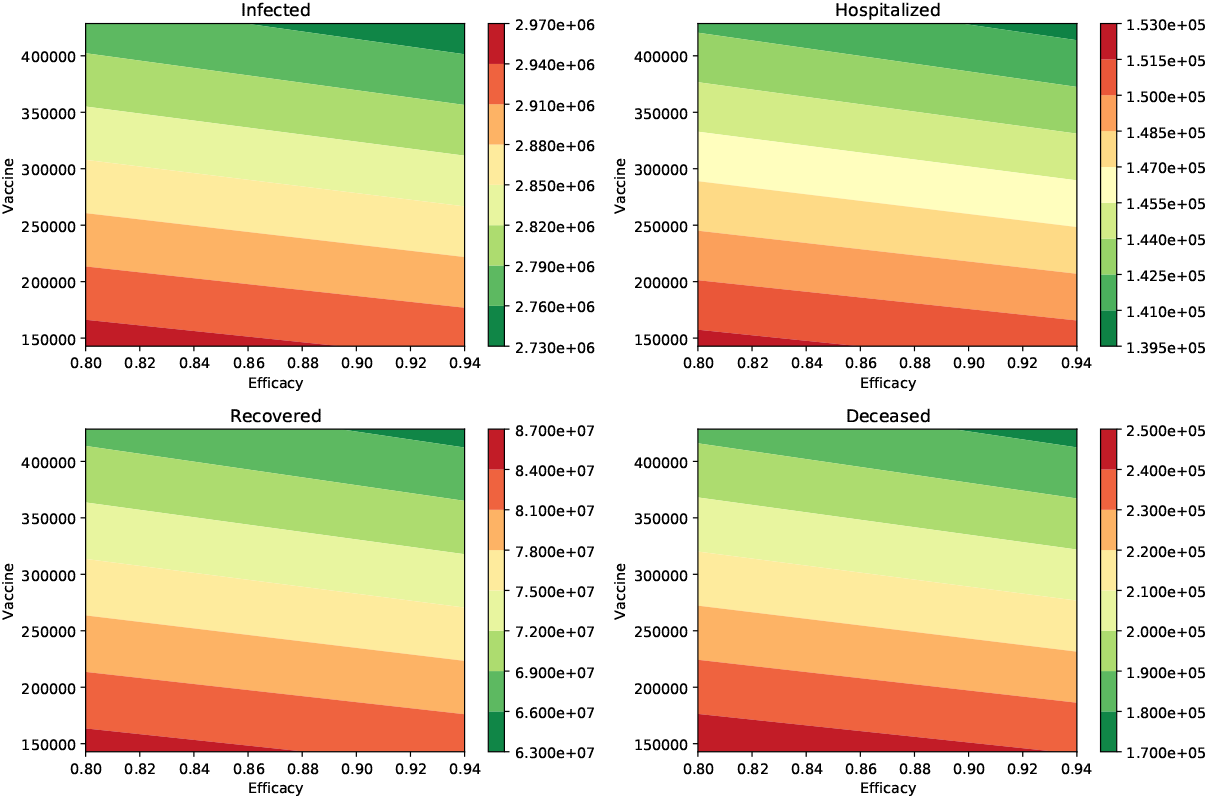
Impact of the inoculation rate and efficacy of the vaccine on the peak of the infected (I) and hospitalized (H) subpopulations. In addition, on the deaths (D) and recovered (R) cases. In these scenarios, the infectiousness of the asymptomatic individuals relative to symptomatic is 75% (β_A_ = β_I_), and the percentage of infections that are asymptomatic is 50%. In all these scenarios the transmission rate is β = 0.2.

Finally, Table 5 and Table 6, show the outcome when the percentage of infections that are asymptomatic is 40% (*a* = 0.4) and the infectiousness of the asymptomatic individuals relative to symptomatic is 100% and 75% respectively. The numerical simulation results show similar trends to the two previously studied cases. Again, that the impact of the vaccination rate is greater than the efficacy of the vaccine can be observed. This qualitative effect can be seen under a variety of scenarios regarding vaccine efficacy and vaccination rate in Figures 4 and 5, respectively. The numerical simulations include many different parameter values for the infectiousness of asymptomatic individuals, percentage of infections that are asymptomatic, efficacy of the vaccine, and the vaccination rate. Thus, uncertainty in these parameters has been considered in this study.

**Table 5:** Impact of the inoculation rate (Vac.) and the efficacy (ϵ) of the vaccine on the peak of the infected (I) and hospitalized (H) subpopulations. In addition, on the deaths (D) and recovered (R) cases. In these scenarios, the infectiousness of the asymptomatic individuals relative to symptomatic is 100% (β_A_ = β_I_), and the percentage of infections that are asymptomatic is 40%.

| Pop | Vac.<br>Eff<br>Trans | 2 millions |  | 4 millions |  |
| --- | --- | --- | --- | --- | --- |
|  |  | 94% | 80% | 94% | 80% |
| I | 0.2 | 5.542887e+06 | 5.637083e+06 | 4.625919e+06 | 4.799400e+06 |
|  | 0.22 | 8.694897e+06 | 8.840743e+06 | 7.180468e+06 | 7.497133e+06 |
| H | 0.2 | 3.426932e+05 | 3.485607e+05 | 2.855083e+05 | 2.963821e+05 |
|  | 0.22 | 5.335221e+05 | 5.425334e+05 | 4.403929e+05 | 4.599537e+05 |
| R | 0.2 | 1.289053e+08 | 1.320051e+08 | 9.968921e+07 | 1.056169e+08 |
|  | 0.22 | 1.624433e+08 | 1.656592e+08 | 1.307745e+08 | 1.380529e+08 |
| D | 0.2 | 4.768158e+05 | 4.897834e+05 | 3.544676e+05 | 3.793021e+05 |
|  | 0.22 | 6.175980e+05 | 6.310708e+05 | 4.848819e+05 | 5.153864e+05 |

**Table 6:** Impact of the inoculation rate (Vac.) and the efficacy (ϵ) of the vaccine on the peak of the infected (I) and hospitalized (H) subpopulations. In addition, on the deaths (D) and recovered (R) cases. In these scenarios, the infectiousness of the asymptomatic individuals relative to symptomatic is 75% (β_A_ = β_I_), and the percentage of infections that are asymptomatic is 40%.

| Pop | Vac.<br>Eff<br>Trans | 2 millions |  | 4 millions |  |
| --- | --- | --- | --- | --- | --- |
|  |  | 94% | 80% | 94% | 80% |
| I | 0.2 | 3.165047e+06 | 3.196961e+06 | 2.886104e+06 | 2.932840e+06 |
|  | 0.22 | 5.103234e+06 | 5.189472e+06 | 4.277550e+06 | 4.430910e+06 |
| H | 0.2 | 1.966936e+05 | 1.987256e+05 | 1.786035e+05 | 1.817093e+05 |
|  | 0.22 | 3.158189e+05 | 3.211994e+05 | 2.642139e+05 | 2.738498e+05 |
| R | 0.2 | 8.946476e+07 | 9.181957e+07 | 6.917128e+07 | 7.274332e+07 |
|  | 0.22 | 1.230787e+08 | 1.261358e+08 | 9.460603e+07 | 1.002532e+08 |
| D | 0.2 | 3.113456e+05 | 3.211641e+05 | 2.264752e+05 | 2.414360e+05 |
|  | 0.22 | 4.523586e+05 | 4.651427e+05 | 3.331442e+05 | 3.568021e+05 |

**Figure 4:**
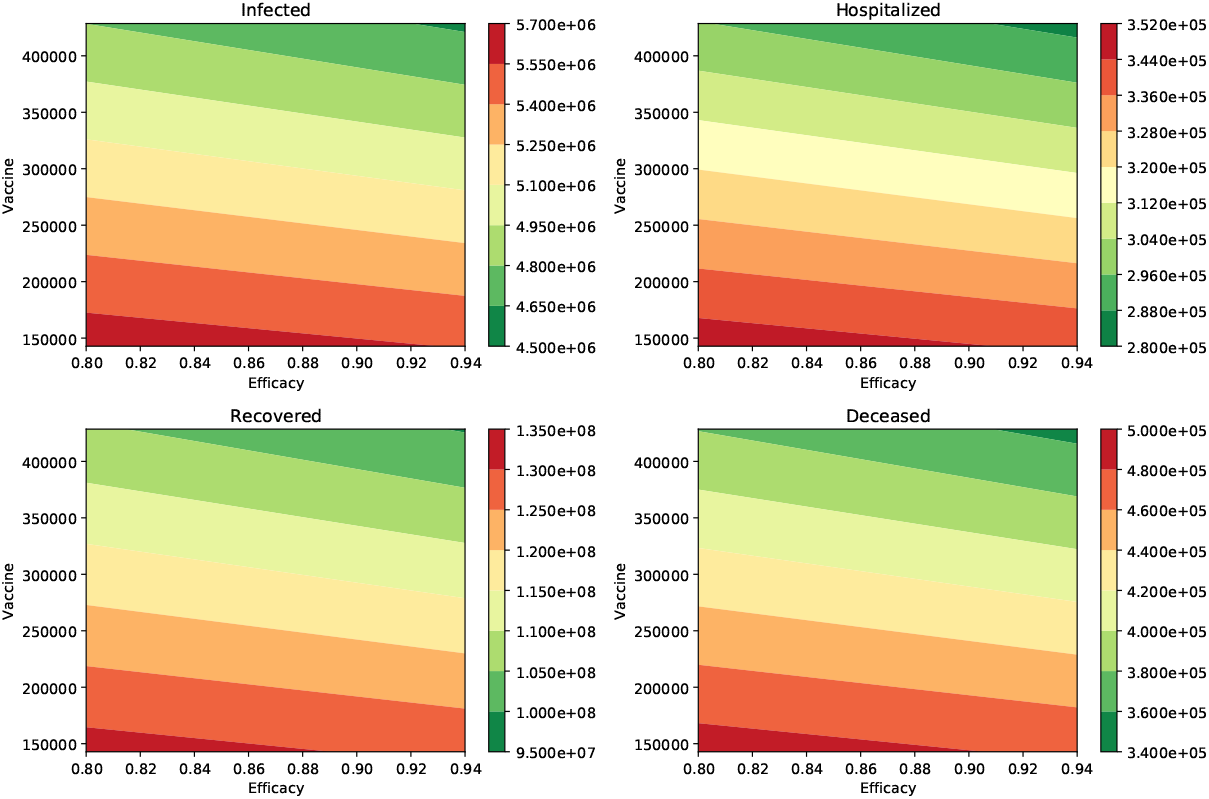
Impact of the inoculation rate and efficacy of the vaccine on the peak of the infected (I) and hospitalized (H) subpopulations. In addition, on the deaths (D) and recovered (R) cases. In these scenarios, the infectiousness of the asymptomatic individuals relative to symptomatic is 100% (β_A_ = β_I_), and the percentage of infections that are asymptomatic is 40%. In all these scenarios the transmission rate is β = 0.2.

**Figure 5:**
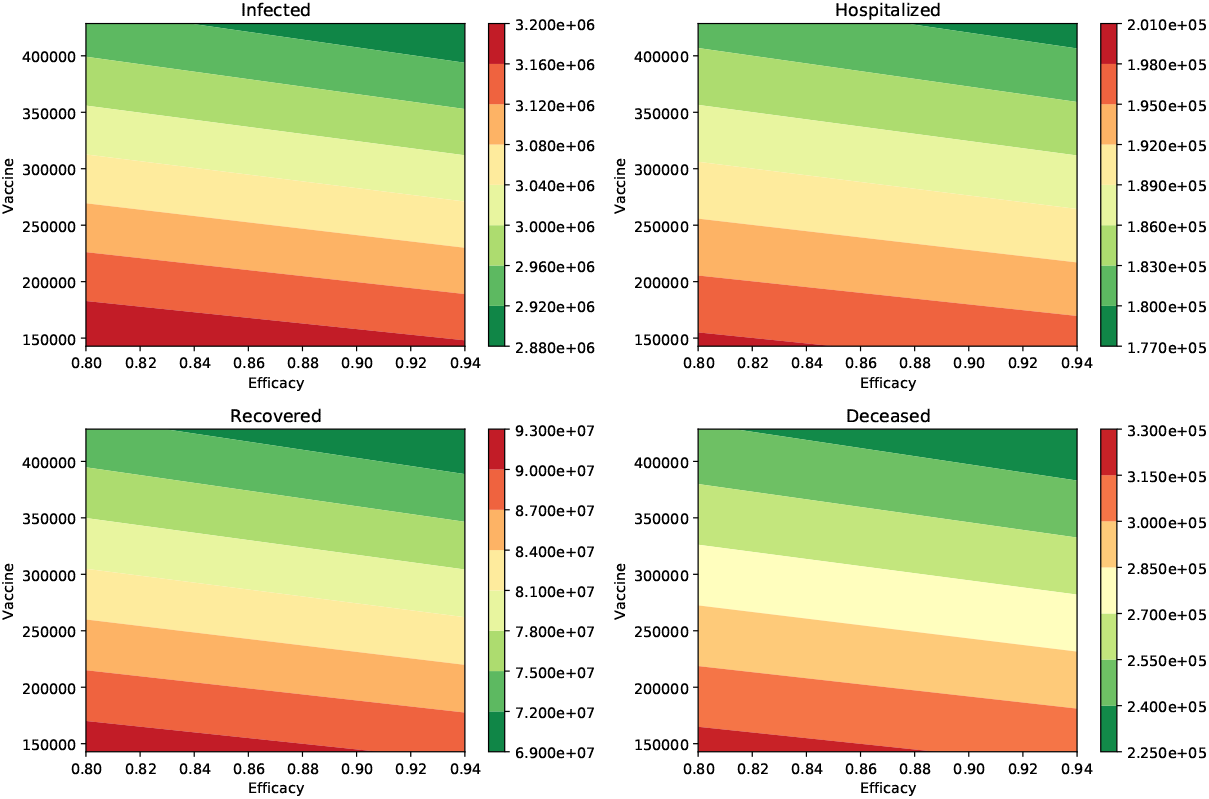
Impact of the inoculation rate and efficacy of the vaccine on the peak of the infected (I) and hospitalized (H) subpopulations. In addition, on the deaths (D) and recovered (R) cases. In these scenarios, the infectiousness of the asymptomatic individuals relative to symptomatic is 75% (β_A_ = β_I_), and the percentage of infections that are asymptomatic is 40%. In all these scenarios the transmission rate is β = 0.2.

Based on the previous results we can conclude that under some plausible scenarios that the impact of the inoculation rate is more relevant to control the burden of the COVID-19 pandemic. Thus, these results suggest that health authorities should focus in increasing the inoculation rate in order to avert more infected people, hospitalizations and deaths. Our results agree with previous result under different assumptions, and with recommendations made by some scholars [99, 141].

## 4 Discussion

Currently there are authorized and recommended vaccines to prevent COVID-19 in the United States. The COVID-19 vaccination program started in early December. Depending on the specific vaccine, the people will get a second shot 3-4 weeks after the first in order to achieve the most protection against the disease caused by the SARS-CoV-2 virus [30, 106]. The vaccines against the SARS-CoV-2 virus have different efficacies and mechanisms of action [3, 16, 69, 71, 149, 105, 135]. Vaccination programs have been recently begun in other countries, using several different types of program and different outcomes can therefore be anticipated [1, 69, 137]. For instance, a vaccination program can focus first on health care workers or on elderly people [87]. However, whatever group the vaccination program targets first; there is an inoculation rate of the vaccine. In this study we propose a mathematical model to assess the impact of the vaccination programs as a function of the efficacy of the vaccine and the inoculation pace. The study of different COVID-19 vaccination programs is of paramount importance to reduce the burden of the COVID-19 pandemic. An optimal vaccination program helps to tackle the transmission of the SARS-CoV-2 virus in the population in an efficient way [9, 29, 69, 72, 137, 145]. It is important to mention that the vaccination programs in different regions or countries vary due to multiple constraints. For instance, there are vaccines that require special storage and transportation, and this affects the availability of the vaccine. Moreover, the current number of vaccine shots are not enough to vaccinate the whole world population [11, 16, 18, 53, 87].

In this article, we studied the impact of the vaccination pace and the efficacy of the vaccine on the dynamics of the COVID-19 pandemic. We studied the particular scenario of USA, but the methodology presented here can be extrapolated to other countries or regions. We were able to study different potential scenarios regarding the burden of the COVID-19 pandemic. We varied the inoculation rate, efficacy of the vaccine and the SARS-CoV-2 virus transmission rates. The constructed compartmental mathematical model allows the variation of the aforementioned factors, and using computational methodologies we obtained metrics that indicate which are the most important factors to decrease the burden of the COVID-19 pandemic.

We found that the efficacy of the vaccine and the vaccine inoculation rate have a high impact on the outcomes. However, the rate of vaccine administration has a larger impact on reducing the infected and hospitalized subpopulations. In a similar way, it has a greater impact on the number of deaths caused by the SARS-CoV-2 virus. Another important finding is that the impact of the inoculation rate and vaccine efficacy is larger for scenarios with higher SARS-CoV-2 virus transmission rates. Thus, our results suggest that health institutions need to focus in increasing the vaccine inoculation rate in the regions with higher rate of new infections. Our results are in accordance with previous recommendations made by some scholars [99, 141].

As expected from a vaccination program the results show that the benefits depend on how it is implemented and the efficacy of the vaccine. As we have mentioned, the total coverage of the population would depend on the production of the vaccine doses and the deployment of resources to execute the vaccination programs. In addition, there is a potential limitation of the total coverage due to the relctance of some parts of the population who have been influenced by doubting the science or by pressure from anti-vaccination groups [27, 53, 108, 137]. Our results also show that the impact of a COVID-19 vaccination program is highly dependent on the SARS-CoV-2 virus transmission rates and these affect the effective reproductive number ℛ_*t*_ of the SARS-CoV-2 virus. Thus, it is important to educate the population about the importance of maintaining non-pharmaceutical control interventions such as the use of facial masks and physical distancing [30, 40, 73, 84, 110].

Furthermore, the additional benefit of a vaccine with 80% or 94% efficacy depends on the SARS-CoV-2 virus transmission rate as has been observed in this study. When we have low SARS-CoV-2 virus transmission rates (equivalently: lower effective reproductive number) the vaccine with 80% efficacy has a smaller impact on the COVID disease related metrics in comparison with a scenario with a high SARS-CoV-2 virus transmission rate (equivalently higher effective reproductive number). Thus, even with a highly effective vaccine it is important to maintain as low as possible the SARS-CoV-2 virus transmission rate to reduce the burden of the current pandemic. Moreover, if the immunity against the SARS-CoV-2 virus diminishes over the time, then lowering transmission rates is even more crucial.

The constructed compartmental model is a SEIR type but with some additional features such the compartment for asymptomatic cases. We expanded the model to include vaccinated people even if the vaccine is not effective in some subset. The SARS-CoV-2 virus spread is mainly driven by the values of the parameters, which have some uncertainty, as is usual in this type of epidemiological model. The uncertainty related to the COVID-19 pandemic is higher in comparison with other diseases such influenza due to the novelty of the SARS-CoV-2 virus. The parameter values were chosen from scientific literature. Despite the limitations of this type of mathematical model, they have been useful in many epidemics and are a classical method to deal with epidemics [57, 22, 28, 45, 34, 13, 36, 73, 12]. Some particular limitations of this study are that constant inoculation rates were used and the vaccination programs do not target any specific subpopulation. We hopefully anticipate that the inoculation rates will increase due to an increase in vaccine production and improving of the logistics. However, the vaccination programs might face several obstacles along the way. The model does not consider a subpopulation that is not willing to take the vaccine, and this has been an issue for other vaccines [27, 53, 108, 137]. Further studies are needed to extend the mathematical model for other vaccination programs. For instance, those that target first health care workers or specific age groups. This would require more parameters and therefore more uncertainty and details. In addition, our mathematical model does not consider the fact that immunity wanes. In fact, the US FDA recommends that follow-up of study participants should continue for as long as is feasible, to assess the duration of protection [58].

## 5 Conclusions

The results presented in this study show that the effectiveness of a COVID-19 vaccination program strongly depends on the vaccination rate and the efficacy of the vaccine. Moreover, the SARS-CoV-2 virus human transmission rates and consequently the effective reproductive number impact the outcome of the vaccination programs. It is important to remark that vaccination rate depends on many variables or resources such as health care facilities or logistical transportation aspects. On the other hand, the efficacy of the vaccine is out of the hands of health institutions and official entities. However, the rate of vaccine administration plays a more important role to reduce the burden of the COVID-19 pandemic. Our results show that health institutions need to focus in increasing the vaccine inoculation pace and create awareness in the population about the importance of the COVID-19 vaccines. In some countries the vaccination rate would be limited due to the availability of the vaccine. Currently, in the USA there are issues with the vaccination rate due to logistics, but not regarding availability [134, 106]. As we mentioned in the introduction, at some point there might be difficulties keeping a constant vaccination rate since a proportion of the population is not willing to be vaccinated. This topic is interesting and can be studied in the future.

The type of mathematical model, based on ordinary differential equations, used here suffers from the following limitations: exponential distributions in the transitions from one stage to another are implicitly assumed as well as homogeneous mixing in the population. Additionally, the behavior of individuals is averaged in order to avoid more complex models that in turn have their own limitations. For instance, individual agent based models have many parameters and in several cases the values of these parameters are very difficult to obtain. However, in some cases exponential transition between stages are not very far from reality. Despite the limitations of our model, we found valuable results to face the current COVID-19 pandemic. Support is given to characteristics of efficient vaccine campaigns. In particular, our study encourages governments and their health institutions to increase the pace of the vaccination in the population in order to diminish the consequences of the catastrophic COVID-19 pandemic.

## Data Availability

Data used in the article is public.

## Notes

### Competing Interest Statement

The authors have declared no competing interest.

### Funding Statement

No external funding.

### Author Declarations

Data is public.

